# OVERWHELMING GENETIC HETEROGENEITY AND EXHAUSTING MOLECULAR DIAGNOSTIC PROCESS IN CHRONIC AND PROGRESSIVE ATAXIAS: *FACING IT UP WITH AN ALGORITHM, A GENE, A PANEL AT A TIME*

**DOI:** 10.1101/2020.03.20.20036996

**Authors:** J Perez Maturo, L Zavala, P Vega, D González-Morón, N Medina, V Salinas, J Rosales, M Córdoba, T Arakaki, N Garretto, S. Rodríguez-Quiroga, MA Kauffman

## Abstract

Ataxias is one of the most frequent complaints in Neurogenetics units worldwide. Currently, more than 50 subtypes of spinocerebellar ataxias and more than 60 recessive ataxias are recognized. We conducted an 11-year prospective, observational, analytical study in order to estimate the frequency of pediatric and adult genetic ataxias in Argentina, to describe the phenotypes of this cohort and evaluate the diagnostic yield of the algorithm used in our unit. We included 334 ataxic patients. Our diagnostic approach was successful in one third of the cohort. A final molecular diagnosis was reached in 113 subjects. This rate is significantly higher in the subgroup of patients with a positive family history, where the diagnostic yield increased to 55%. The most prevalent dominant and recessive ataxias in Argentina were SCA-2 (36% of dominant ataxias) and FA (62% of recessive ataxias), respectively. Next generation sequencing based assays were diagnostic in the 65% of the patients requiring these tests. These results provide relevant epidemiological information, bringing a comprehensive knowledge of the most prevalent subtypes of genetic ataxias and their phenotypes in our territory and laying the groundwork for rationally implementing genetic diagnostic programs for these disorders in our country.

## 1. INTRODUCTION

Ataxias is one of the most frequent complaints in Neurogenetics units worldwide. The global prevalence of autosomal dominant (AD) ataxias is estimated to be 1 to 5/100,000 inhabitants. Among them, Spinocerebellar Ataxia type 3 (SCA-3) is the most prevalent. The worldwide prevalence of recessive ataxias (RA) is similar, being Friedreich’s Ataxia (FA) by large the most common in this group [1-3]. Currently, more than 50 subtypes of spinocerebellar ataxias and more than 60 recessive ataxias are recognized [3-6]. Their overwhelming genetic heterogeneity is one of the most representative features of this group of disorders.

The molecular diagnostic process in this field is often exhausting, since it frequently involves the request and execution of numerous tests with the aim of identifying the etiopathogenic genetic defect. Moreover, diverse mutational mechanisms (such as point mutations, structural anomalies and trinucleotide repeat expansions) can be present, requiring specific techniques to individualize them. In addition, considering that most of the molecular tests are expensive and many are not easily accessible in less developed countries, it is fundamental to apply a rational diagnostic methodology that maximize the probability of a positive result in each individual patient. For this, it is necessary to know the frequency of the different ataxias in the population under study and the distinctive phenotypes of each of them.

In spite of isolated case reports and descriptions of small series of patients with inherited ataxia [7-11] and preliminary results presented by our group [12], there has not been previous report of a systematic analysis that has evaluated the diagnostic performance of a comprehensive molecular diagnostic approach for these group of genetic disorders in Argentina. We conducted an 11-year prospective, observational, analytical study in order to estimate the frequency of pediatric and adult genetic ataxias in Argentina, to describe the phenotypes of this cohort and evaluate the diagnostic yield of the algorithm used in our unit.

## 2. PATIENTS AND METHODS

### 2.1. Patients

A prospective, observational, descriptive and analytical study of adult and pediatric patients with a progressive ataxia was carried out. We included all patients that have been evaluated between May 2008 and December 2019 within the *Chronic and Progressive Ataxia Program* that is held at the Neurogenetics Unit and Movement Disorders Area from the Neurology Division of the Hospital J. M. Ramos Mejia-Buenos Aires, Argentina. All adult patients and parents of pediatric patients gave their informed consent to participate and the study was approved by the institutional ethics committee of our institution. In all of the patients we used a structured clinical interview in order to register demographic characteristics, family history and the main clinical features of their complain. The semiquantitative SARA scale was used to evaluate cerebellar dysfunction and quantification of the severity of ataxia. This scale has been previously validated to stratify severity in patients with SCAs and FA. [13, 14].

The clinical examination included the exploration of horizontal and vertical saccadic and smooth pursuit eye movements, absence or presence of nystagmus, and signs of oculomotor apraxia. The presence/absence of movement disorders such as chorea, myoclonus, dystonia, parkinsonism or tremor was also explored. Peripheral neuropathy was defined as the presence of a decrease or absence of osteotendinous reflexes, superficial or deep sensory distal involvement, and/or electrophysiological evidence of nerve conduction abnormalities. The presence of hyperreflexia, Hoffman signs and/or extensor plantar reflexes were recognized as signs of pyramidal dysfunction. The presence of cerebellar atrophy was evaluated by magnetic resonance brain images. Routine laboratory was analyzed and biochemical markers such as serum alpha-fetoprotein, albumin, electrophoretic proteinogram, metabolic profile and creatine phosphokinase (CPK) were determined.

### 2.2. Patient stratification. Selection of Genetic Diagnostic Approach

The patients were stratified according to their family history, the age at disease onset and the phenotypic features than can be considered distinctive/suggestive of particular etiologies **(Fig. 1a, 1b, 1c)**.

**Figure 1A.**
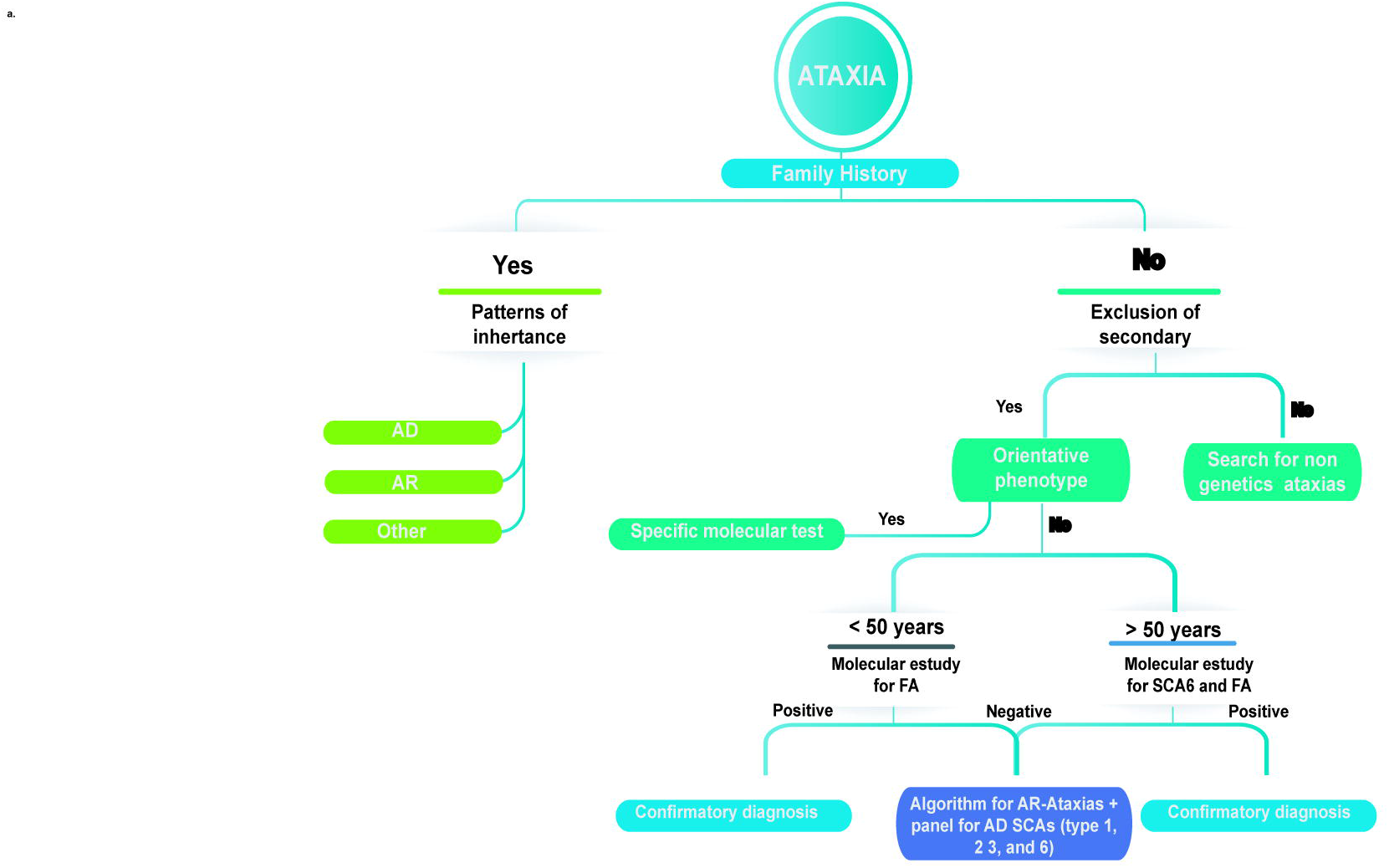
Initial diagnostic approach. **AD**: Autosomal Dominant; **AR**: Autosomal Recessive; **FA:** Friedreich’s Ataxia; **SCA**: Spinocerebellar Ataxia

**Figure 1B.**
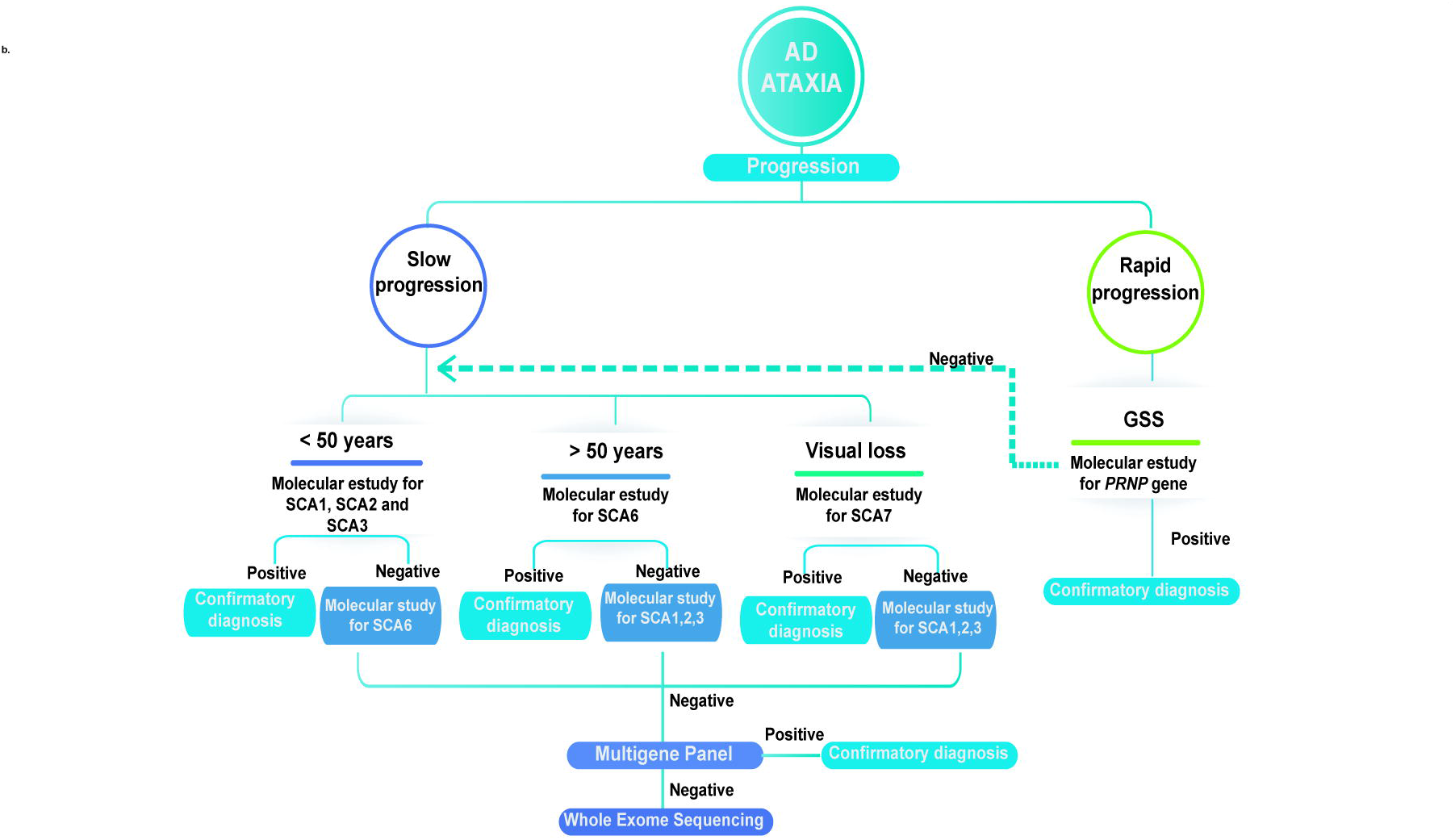
Diagnostic approach for Autosomal Dominant Ataxias. **AD**: Autosomal Dominant; **SCA**: Spinocerebellar Ataxia; **GSS**: Gerstmann-Sträussler Scheinker disease

**Figure 1C.**
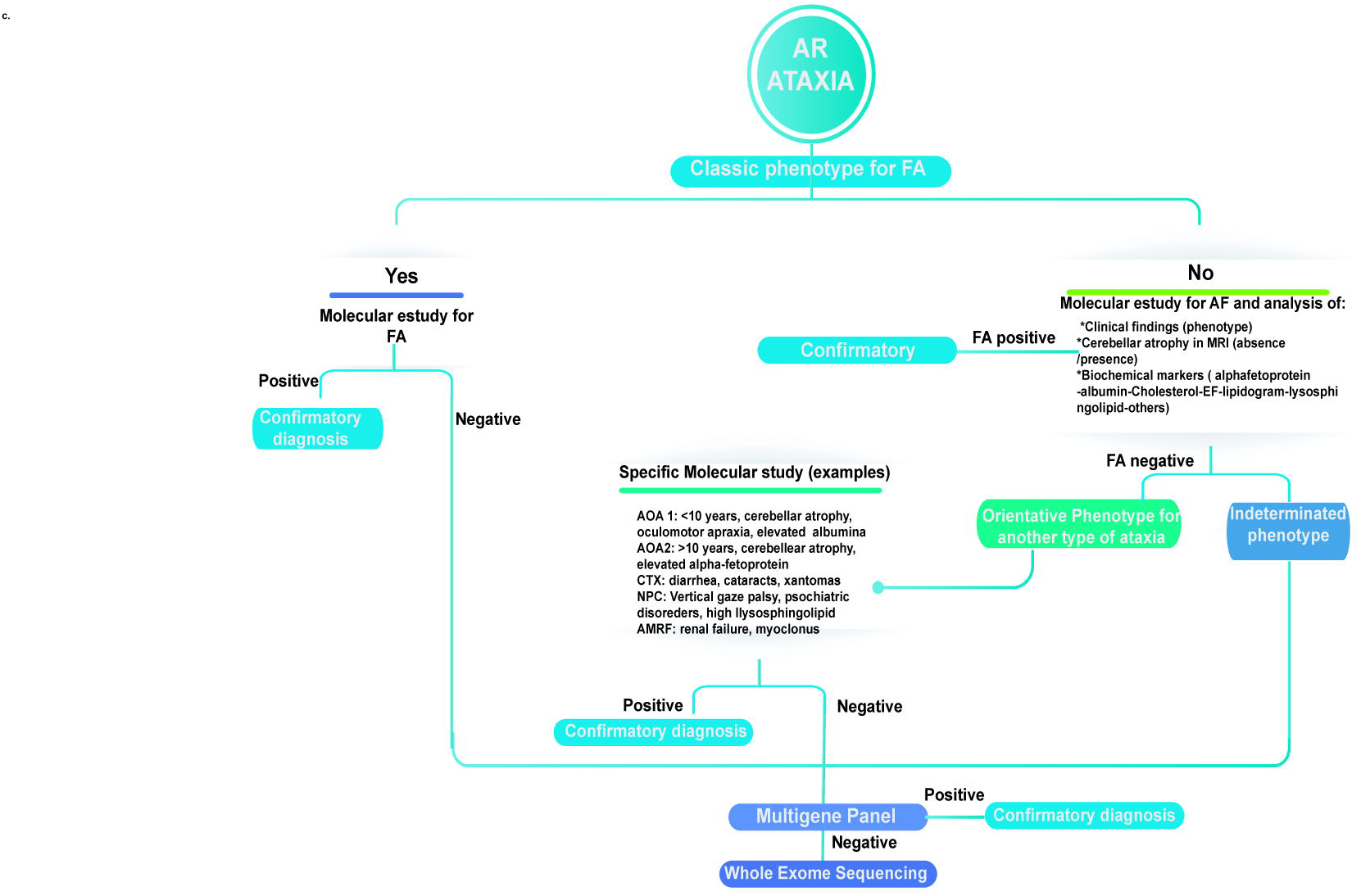
Diagnostic approach for autosomal Recessive Ataxias. **AR**: Autosomal Recessive; **FA:** Friedreich’s Ataxia; **AOA:** Ataxia with oculomotor apraxia **; CTX:** Cerebrotendinous xanthomatosis; **NPC:** Niemann Pick-C disease; **AMRF** Action myoclonus–renal failure syndrome

#### 2.2.1. Autosomal dominant Ataxias (AD)

In all patients with a pedigree consistent with an autosomal dominant inheritance pattern, we performed a multigene panel of AD ataxias that included tests for the detection of abnormal repeat expansions of CAG causing the most prevalent SCAs subtypes worldwide (SCA-1, SCA-2, SCA-3 and SCA-6). The phenotype of each individual patient set the order that each of these tests was performed. In cases where the phenotype was not distinctive of a specific form of SCA, the complete panel of dominant ataxias was performed simultaneously. If the phenotype was characteristic of a less prevalent SCA or other dominant Ataxia, specific molecular studies were performed based on our clinical suspicion (e.g. SCA-7, Huntington’s disease, Gerstmann-Straussler-Scheinker disease among others) **(Fig 1b)**.

#### 2.2.2. Autosomal recessive Ataxias (AR)

In the group of patients with an autosomal-recessive inheritance, we looked first for abnormal expansions of GAA repeats in the intron 1 of *FXN* gene. In case of a negative result, we took into account the phenotype, biochemical markers and neuroimages to define if the patients met features suggestive of a specific disease (e.g. Niemann Pick type C disease, Ataxia with Oculomotor Apraxia 1 or 2, Cerebrotendinous Xanthomatosis among others) in order to sequence individual genes. In cases where the subjects presented undetermined phenotypes, we used next generation sequencing assays (described in next sections). **(Fig. 1c)**

#### 2.2.3. Ataxia without the antecedent of affected relatives (sporadic cases)

For sporadic ataxia patients, in whom secondary causes were ruled out, we took into account the phenotype, biochemical markers and neuroimages to define if the patients met features suggestive of a specific disease that would direct us for the sequencing of individual genes. If they presented with undetermined phenotypes, we considered their age at onset: in subjects with an age at onset below 50 years, tests for FA were done first, otherwise, tests for FA and SCA-6 were done at the same time (Figure 1a). Finally, when a confirmatory diagnosis was not obtained after these initial tests, the diagnostic approach continued as in AD and AR ataxias simultaneously (described in previous sections).

#### 2.2.4. Ataxias with non-diagnostic first tier tests. Next Generation Sequencing Assays

Finally, those cases with non-diagnostic first tier tests were studied by means of NGS assays that included multigene panels, exome sequencing or genome sequencing according to availability in each case **(Fig**.**1b, 1c)**.

### 2.3. Genetic Diagnostic Procedures

Two millimeters of whole blood were obtained from each patient by venipuncture. Total genomic DNA was purified using commercial systems following the manufacturer’s instructions. Evaluation of trinucleotide repeats was performed by PCR amplification with gene-specific fluorescent labelled primers and fragment length analysis following capillary electrophoresis according to standard procedures previously by our group [12]. For FA diagnosis, we performed fluorescent PCR-electrophoresis fragment sizing, Triplet Repeat Primed-PCR (TP-PCR) and eventually complete *FXN* coding regions sequencing by Sanger method as previously described [12, 15]. Similarly, Sanger sequencing-based assays were used for the analyses of *PRNP, NPC1, NPC2, CYP27A1, SCARB2* genes. Next generation sequencing assays were performed according to previous methods described by our group, TruSight One (Illumina INC) based multigene panel sequencing in [16] and exome sequencing in [17]. Bioinformatics procedures and variants interpretation was done as previously described by our group in [16, 17].

## 3. RESULTS

### 3.1. Cohort. Global Analysis

A consecutive population of 1250 patients were evaluated between May 2008 and December 2019 at the Neurogenetics Unit of a tertiary neurology service in a public hospital in Buenos Aires, Argentina. We prospectively included 334 subjects for this study because they presented with a chronic cerebellar-ataxic dysfunction of presumed genetic etiology. **Table 1** presents a summary of the demographic characteristics of this population. We systematized the study of these patients following the algorithm presented in **Fig. 1**, selecting the type and order of molecular genetic tests in each case. The average age of our cohort was 41 years, involving a wide range of ages that spanned from children to the elderly (range 2-83 years). A similar number of females and males attended our clinic, with a slight predominance of female sex (F: 56.5% / M: 43.5%). Nearly 46% of the patients (154 subjects) had a positive family history for a similar condition. The mean age onset was 31.2 years (range <1-81 years). The so-called *“diagnostic odyssey”* in the subgroup of patients with a positive molecular diagnosis extended for an average lapse of 10.07 years (range 0-60 years).

**TABLE 1:** Demographic Characteristics of the general population.

| Demographic Characteristics |  |
| --- | --- |
| Total number of patients with ataxia | 334 |
| Gender |  |
| Male (%) | 56.5% |
| Female (%) | 43.5% |
| Mean age in years (min-max) | 41 (2-83) |
| Positive family History (%) | 46% |
| Inheritance patterns (%) |  |
| AD | 73% |
| AR | 27% |
| Mean Age at onset (min-max) | 31.2 (<1 -81) |
| Diagnostic delay in years (min-max) | 10.07 (0-60) |

AD: Autosomal Dominant; AR: Autosomal Recessive

The algorithm implemented was successful in one third of the patients (33,8%). A final molecular diagnosis was reached in 113 subjects. This rate is significantly higher in the subgroup of patients with a positive family history, where the diagnostic yield increased to 55%. In those with an AD inheritance, a molecular diagnosis was obtained in 41 of 81 families (50,6%), whereas in cases where the family history was suggestive of recessive inheritance, the diagnosis was reached in 20 of 29 families (69%) **(Table 2)**.

**TABLE2.** Pedigrees with confirmatory molecular diagnoses.

|  |  |
| --- | --- |
| N of pedigrees with positive family history | 110 families |
| Confirmatory molecular diagnosis | 55% [61/110] |
| N of pedigrees with positive Autosomal Dominant Inheritance | 41 families |
| Confirmatory molecular diagnosis | 50,6% [41/81] |
| N of pedigrees with positive Autosomal Recessive inheritance | 20 families |
| Confirmatory molecular diagnosis | 69% [20/29] |
| Patients with Sporadic Ataxia | 180 patients |
| Confirmatory Molecular Diagnosis | 16% [29/179] |

N: number

### 3.2. Ataxias with identified genetic etiology

In the following subsections, the clinical and molecular details of the 113 patients in whom a genetic etiology was identified. The cohort is divided according to the inheritance mechanism and the requirement for diagnosis of Next Generation Sequencing tests.

#### 3.2.1. Autosomal Dominant Ataxias

55 patients were diagnosed with a form of AD ataxia. The most frequent was SCA-2 (36%), followed by SCA-3 (18%) and SCA-1 (145%). A detail of the rest of AD ataxias is presented in **Supplementary Material-Fig. S1a and Table S1**. The distribution by gender was slightly predominant in women (F: 54.5% / M: 45.5%) and the average age at first consultation was 40,1 years (range 2-69 years). The mean age at onset was 30.7 years (range <1-63 years), whereas the mean time until a molecular diagnosis was 9.1 years. With the exception of two sporadic patients, diagnoses of SCA-2 and KCNA2 early onset ataxia, all of the subjects of this group have the antecedent of multiple affected members in their family.

We observed a wide range of severity of cerebellar dysfunction according to the SARA scale (0-32 points). A rapidly progressive course and severe cerebellar involvement was characteristic of the four subjects that received a diagnosis of GSS. The presence of markedly slow horizontal saccades was typical of SCA-2, we found it in 15 of the 20 (75 %) subjects that received this diagnosis. A summary of eye movements abnormalities in the cohort is presented in **Supplementary Material-Table S2**. A number of different extra-cerebellar abnormalities was found in these patients **(Supplementary Material-Table S3)**. Parkinsonism and polyneuropathy were the most frequent extra-cerebellar findings in SCA-3 and SCA-2 patients. Myoclonus was found in two patients with SCA-2, in one with SCA-3 and in one of the subjects with GSS. Pyramidal tract involvement was common in SCA-1 and in SCA-2 patients. Cognitive impairment was infrequent, except in one subject with GSS and in another with a molecular diagnosis of HD that showed an atypical ataxic dominant presentation. Cerebellar atrophy was identified in the vast majority of MRIs of this group (76%).

#### 3.2.2. Autosomal Recessive Ataxias

58 subjects were diagnosed with a form of autosomal recessive ataxia **(Supplementary Material-Table S4)**. FA was largely the most frequent subtype. It accounted for a 62% (36/58) of the diagnoses of this group. A detail of the rest of recessive ataxias patients diagnosed here is presented in **Supplementary Material-Fig. S1b and Table S4**. The distribution by gender showed an excess of male patients (M: 58.4% / F: 41.6%). The mean age at consultation was 27 years (range 4-75 years). The mean age at onset of symptoms was 13.2 years (range 1-33 years) with a delay since the first symptom until the final diagnosis of about 15 years. About half of the subjects (27/58) had a sporadic presentation.

The vast majority of FA patients manifested a classical phenotype (31/36). We identified five subjects with late onset FA **(Supplementary Material-Table S5)**. We observed a mean SARA scale of 20 (range 6-39 points). Eye movement abnormalities were common in FA patients. We observed macro-square waves (15/36) and hypometric horizontal (20/36) and vertical saccades (19/36) in the majority of FA patients **(Supplementary Material-Table S6)**. Among the extra-neurological findings of this group of patients, we can mention the presence of myocardiopathy (8/36), pes cavus (8/36) and diabetes mellitus (1/36) **(} Supplementary Material-Table S5)**. With the exception of 1 patient that was compound heterozygous for a missense variant and an intron 1 expanded allele, all of the FA patients were compound heterozygous for abnormally expanded alleles in the intron 1 of FXN gene. The description of the rest of subjects with other forms of RA is shown in **Supplementary Material-Table S4**.

#### 3.2.3. Next Generation Sequencing based Ataxia Diagnoses

In a subset of 26 patients, we needed to use next generation sequencing based assays (multigene panels, exome sequencing and whole genome sequencing) for diagnostic purposes **Supplementary Material-Table S7**. This approach proved to be successful in more than half of the cases (17/26). A multigene panel was used in 14 subjects, arriving to a final diagnosis in 10 of them. A 51-years-old patient consulting for a longstanding familial ataxia was diagnosed with SCA-28 after the identification of a pathogenic variant in *AFG3L2* gene. A diagnosis of AOA type 2 was done in two patients, one is a male patient affected with a progressive sporadic ataxia and the other, is a female patient with oculomotor apraxia, cerebellar atrophy and elevated alpha-fetoprotein serum level. Furthermore, four early onset ataxia cases were resolved after finding pathogenic variants in *CC2D2A* and *ATM* genes (3 cases), respectively. In addition, a patient was diagnosed with a cerebellar phenotype of a late infantile neuronal ceroid lipofuscinosis due to pathogenic variants in *TPP1* gene; a 37–years-old man with a pure cerebellar syndrome was diagnosed as ARCA-1 disease due the presence of two pathogenic variants in the *SYNE1* gene, and two female patients with ataxia and supranuclear gaze palsy received a diagnosis of NP-C. Exome sequencing was performed in 11 subjects, this test proved to be useful in 6 patients. An 8-year-old patient that showed progressive ataxia, chorea and cerebellar atrophy received a diagnosis of AOA type 1 after finding pathogenic variants in *APTX*. In another patient showing a very early onset ataxia accompanied with a seizure disorder, a diagnosis KCNA2-related encephalopathy was done. Moreover, three other patients received a diagnosis after finding pathogenic variants in *STUB1*(2 cases), *OPA1 a*nd *SACS* genes, respectively. Finally, whole genome sequencing was used for the diagnosis of a family showing a complex phenotype characterized by the presence of episodic ataxia and epileptic encephalopathy. This approach allowed us to identify a pathogenic variant in *SCN2A*.

## 4. DISCUSSION

In this study, a pediatric and adult population of hereditary ataxia patients was characterized, obtaining a genetic diagnosis in a significant number of individuals from this cohort. We were able to correctly identify the causing genetic defect in about one third of the patients that were included in our program. We highlight the use of a diagnostic process that rest in a sequential approach directed by the presumptions that arise after individualized and thorough phenotypic characterization of each case. This academic-based approach proved to be useful in terms of diagnostic yield and economy of resources. To the best of our knowledge, the series presented here is the largest ever reported in our country. These results provide relevant epidemiological information, bringing a more comprehensive and precise knowledge of the most prevalent subtypes of genetic ataxias and their phenotypes in our territory. This information has utility for a more rational use of the available resources for the genetic diagnostic approach of ataxic patients. Furthermore, we present evidence of the usefulness of next generation sequencing based assays in patients with uncharacteristic or complex phenotypes or when the most frequent subtypes of hereditary ataxias were previously ruled out. In addition to their high diagnostic yield, their use often avoids unnecessary nonspecific tests with the benefit of reducing costs and shortening the so-called “*Diagnostic Odyssey*” [18-20] for these complex patients.

In relation to similar experiences in other countries, we found similarities in terms of diagnostic yield and techniques used. Brusco et al studied 225 ataxic patients obtaining a genetic diagnosis in 49% of them [21]. A multicenter study in the United States and Canada showed that in 361 families with ataxia, a genetic cause could be identified in 62% of them [22]. A recent study that included 864 ataxic patients showed a diagnostic yield of 30.8% [23]. We found a higher prevalence of SCA-2 than what has been previously reported in other regions where SCA-3 has been the most frequent one [24-26]. However, it is well known that in some countries, particular SCAs are more prevalent, such as the case of SCA-2 in Cuba and SCA-10 in Mexico [27]. Nevertheless, the global prevalence of SCA-2 and SCA-3 found in our series is close to previously reported estimates in western countries [28, 29]. This might be expected considering the important migratory flow from European countries that Argentina has received during the last two centuries. Friedreich ataxia is the most common recessive ataxia worldwide [2, 30, 31]. Unsurprisingly, FA was often found in our cohort as well. It represented about two-thirds of our recessive ataxia diagnoses.

SCAs often compromise extra-cerebellar structures, resulting in florid phenotypes that in some occasions show distinctive features that may direct the genetic diagnostic approach. The presence of unusually slow horizontal saccades in SCA-2 is one of these distinctive flags [32-34]. Accordingly, we observed the presence of markedly slow horizontal saccades in 80% of our SCA-2 cases. Although FA patients often show a classic phenotype at early ages of onset, atypical findings can be observed in later onset cases such as what is reported here for five of our patients. The opposite can be said for non-FA recessive ataxias, where complex and atypical phenotypes are the rule and large genetic heterogeneity is present in every cohort. Accordingly, we found abnormalities in 12 different genes in our cohort of non-FA recessive ataxias. Moreover, multigene next generation sequencing based assays proved particularly useful for the diagnosis of these patients in line with previous reports [35].

## 5. CONCLUSION

In conclusion, we were able to successfully combine the work of clinical and academic based laboratory resources of a tertiary hospital, offering a comprehensive picture of the mutational landscape of genetic ataxias in Argentina, laying the groundwork for rationally implementing genetic diagnostic programs for these disorders in our country. Nevertheless, certainly more work is still needed to further improve the diagnosis in this rapidly evolving field, with the ultimate goal of reducing the individual and familial impact of these devastating neurodegenerative conditions.

## Data Availability

ALL DATA IS AVAILABLE AT THE MANUSCRIPT

## DECLARATIONS

### Disclosure of potential conflicts of interest

#### Ethics approval

This study was approved by the Institutional Ethics Committee of the Hospital JM Ramos Mejia of Buenos Aires, Argentina. All patients and parents provided written informed consent for genetic analyses and use of their anonymized data. All experiments and methods were carried out in accordance with the relevant guidelines and regulations of the Institutional Ethics Committee of the Hospital JM Ramos Mejia of Buenos Aires, Argentina. All clinical investigations have been conducted in accordance with the 1964 Helsinki Declaration and its later amendments or comparable ethical standards.

#### Consent to participate

Informed consent for genetic analyses and use of their anonymized data, was obtained from all individual participants and/or parents included in the study.

#### Consent to publish

Patients signed informed consent regarding publishing their data

## 6. ACKNOWLEDGEMENTS

We thank the patients and families for their support and collaboration.

